# Perioperative outcomes and platinum resistant recurrence in patients undergoing protocol based total parietal peritonectomy during interval cytoreductive surgery for advanced ovarian cancer- results of the TORPEDO study

**DOI:** 10.1101/2023.10.08.23296709

**Authors:** Aditi Bhatt, Snita Sinukumar, Dileep Damodaran, Ankita Patel, Sakina Shaikh, Loma Parikh, Gaurav Goswami, Anjali Singh, Praveen Kammar, Sanket Mehta

**Affiliations:** Dept of Surgical Oncology, Zydus hospital, Ahmedabad, India; Dept of Surgical Oncology, Jehangir Hospital, Pune, India; Dept of Surgical Oncology, MVR cancer center, Calicut, India; Dept of Pathology, Zydus hospital, Ahmedabad, India; Dept of Radiology, Zydus hospital, Ahmedabad, India; Dept of Surgical Oncology, KD hospital, Ahmedabad, India; Dept of Surgical Oncology, Saifee Hospital, Mumbai, India

**Keywords:** Interval cytoreductive surgery, Total parietal peritonectomy, advanced ovarian cancer, HIPEC, platinum resistant recurrence

## Abstract

**Background and aim:** The TORPEDO (CTRI/2018/12/016789) is the single-arm, prospective, interventional study evaluating the role of a total parietal peritonectomy (TPP) in patients undergoing interval cytoreductive surgery (iCRS). In this manuscript, we report the perioperative outcomes and platinum resistant recurrence (PRR) in 218 patients enrolled in the study.

**Methods:** A TPP was performed in all patients undergoing iCRS irrespective of the residual disease extent. HIPEC was performed as per the clinician’s discretion with 75mg/m2 of Cisplatin. Maintenance therapy was also used at the discretion of the treating clinicians.

**Results:** From 9^th^ December 2018 to 31^st^ July 2022 (recruitment complete), 218 patients were enrolled at 4 Indian centres. The median surgical PCI was 14 and a complete gross resection was achieved in 95.8%. HIPEC was performed in 130 (59.6%) patients. The 90-day major morbidity was 17.4% and 2.7% patients died within 90 days of surgery. Adjuvant chemotherapy was delayed beyond 6 weeks in 7.3%. At a median follow-up of 19m [95% CI 15-35m], 101 (46.3%) recurrences and 19 (8.7%) deaths had occurred. The median PFS was 22m [95% CI 17-35m] and the median OS not reached. Platinum resistant recurrence was observed in 6.4%. The projected 3-year OS was 81.5% and in 80 patients treated before may 2020, it was 77.5%.

**Conclusions:** The morbidity and mortality of TPP with or without HIPEC performed during iCRS is acceptable. The incidence was of PRR is low. Early survival results are encouraging and warrant conduction of a randomized controlled trial comparing TPP with conventional surgery.

## Introduction

Patients with advanced ovarian cancer (Stages IIIC and IV) are often treated with neoadjuvant chemotherapy (NACT) followed by interval cytoreductive surgery (iCRS) with three randomized trials reporting non-inferiority to primary cytoreductive surgery (pCRS) with increased rates of complete resection and reduced morbidity. [1,2,3] The survival results, nevertheless, are modest with the median OS in these trials ranging from 29-44 months and the PFS from 10-14 months. [1,2,3] Large retrospective cohort studies from centers of expertise have reported similar outcomes.[4,5] The goal of iCRS is to obtain a complete gross resection or complete cytoreduction (CC-0 resection) which is achieved by resecting sites of visible residual disease.[6] The peritoneal resection that is performed during iCRS could be termed ‘selective parietal peritonectomy (SPP)’.[6] In advanced ovarian cancer, after first line treatment, approximately 10% of the patients will never develop a recurrence.[7] For the remaining 90%, the disease will invariably recur, with the peritoneum being the only or one of the sites of recurrence in more than 80% of the patients.[8] In stages III-C and IV, the diaphragmatic peritoneum is invariably involved.[9] It has been shown that despite having a normal appearance after NACT, microscopic residual disease is present in 15-35% of the patients.[10,11] In other solid tumors, like breast cancer, oral cancers etc., when NACT is used, the resection margin that should be considered (pre chemotherapy versus post chemotherapy) is a topic of debate.[12,13] We hypothesized that removal of the entire parietal peritoneum after NACT could reduce the incidence of peritoneal recurrence and prolong the progression-free and overall survival. The TORPEDO (total removal of the peritoneum at interval debulking for ovarian cancer) study evaluating the role of a total parietal peritonectomy (TPP) during iCRS was initiated in 2018 at three Indian centres. The fourth centre joined the study in September 2020. The perioperative outcomes in the first fifty patients and the incidence platinum resistant recurrence in the first seventy patients enrolled in the study have already been published.[14,15] In addition, the pathological findings in the first 50 patients provided the proof-of-principle for performing a TPP during iCRS.[16] The study completed recruitment in July 2022. In this manuscript, we report the perioperative outcomes and early recurrence in 218 patients enrolled in the study.

## Methods

TORPEDO is a prospective, interventional, multi-centre study, registered with the clinical trials registry of India (CTRI/2018/12/016789). The study was approved by the Zydus Hospital Ethics committee on 8 December 2018. Subsequently, approval was obtained at the other participating institutions.

### Study Protocol

The study enrolled patients with stage III-C or IV-A serous epithelial ovarian cancer undergoing iCRS after NACT. There were not predictive scores used to select patients for pCRS or iCRS but the decision making was individualized. Patients in whom a CC-0/1 resection or complete gross resection (CGR) was not possible, those with a poor performance status or severe ascites were treated by NACT followed by iCRS at all the centres. Patients with stage IV-A disease and negative pleural fluid cytology after NACT were included. The study excluded patients with non-serous histologies, those refusing consent, and those with complete bowel obstruction and/or a poor performance status (Eastern Cooperative Oncology Group [ECOG]>1).

An interim evaluation was performed after three cycles of NACT. Patients with more than a 30% reduction in maximum tumour diameter on imaging and/or whom the surgeon deemed resectable were scheduled for interval CRS. Surgery was performed 3–4 weeks after completion of the third cycle. Administration of more than three cycles was at the discretion of the treating physicians and usually was done to obtain a sufficient response enabling a complete cytoreduction or for logistic concerns.

The surgical protocol has been published previously. Since publication of the OVHIPEC1 trial, HIPEC in the interval setting has been introduced in the National Comprehensive Cancer Network (NCCN) guidelines, which are widely followed in India.[6,17] However, because HIPEC requires an out-of-pocket expenditure for patients, it was performed only for those patients who could afford the additional cost and were willing to undergo the procedure.

HIPEC was performed with cisplatin 75 mg/m^2^ for 90 min by the open (3 centres) or closed (1 centre) method. Adjuvant chemotherapy was initiated for all the patients within 4–6 weeks after surgery. Maintenance therapy was at the discretion of the treating physician. A quality-of-life analysis was to be performed before surgery and at 3, 6, 9, and 12 months after surgery for all the patients.

### Study End Points and Sample Size

The primary end point of the study was progression-free survival, and the secondary end points overall survival, major morbidity, quality-of-life outcomes, and the impact of chemotherapy response grade on survival. A median progression-free survival of 15–18 months is obtained with standard surgery (SPP). We anticipated a 30% benefit in PFS with TPP. To detect this difference with a power of 80% and an alpha of 0.05, 150 patients are required. We aimed to recruit 200 patients to account for some loss of follow-up that could occur.

### Evaluation of Morbidity

This 90-day morbidity and mortality after TPP as part of interval CRS were recorded for all patients. The common toxicology criteria for adverse events (CTCAE) version 4.3 classification was used to record morbidity.[18] Grades 3 and 4 complications were considered major morbidity. Deaths occurring due to any cause were included in the 90-day mortality.

### Surgical Protocol

The goal of surgery was to obtain a complete cytoreduction (CC-0 resection) or CGR.[19] Briefly, a midline incision from the xiphoid to the pubis was performed in all cases irrespective of disease extent. The disease was quantified using Sugarbaker’s Peritoneal Cancer Index (PCI) which was recorded as the surgical PCI (sPCI).[18] TPP comprised all five peritonectomies (pelvic, bilateral antero-parietal, right and left upper quadrant peritonectomies with a total omentectomy). The surgical protocol, regions to be resected for each peritonectomy and the anatomic boundaries have been described and defined in **Supplement 1**.

For mesenteric disease, performance of mesenteric peritonectomy was at the discretion of the surgeon. A minimal representative biopsy of tumour-bearing or normal mesenteric peritoneum (for patients with no ‘visible’ disease) was performed separately for each of the regions from 9 to 12.

Any viscera infiltrated by the tumour were resected to obtain a complete cytoreduction. Visceral sites showing a complete clinical response were not resected the same as the parietal peritoneum. However, some areas of visceral peritoneum were removed if thickened (e.g., reflection of peritoneum from the paracolic gutter over the right colonic mesentery and the posterior layer of the greater omentum reflecting onto the stomach). The pancreatic capsule and Glisson’s capsule were removed only when they had visible disease.

A bilateral pelvic and retroperitoneal lymphadenectomy was performed for all patients with suspicious lymph nodes on imaging or intraoperatively. The upper extent was at the level of the renal veins. In case of suprarenal lymphadenopathy, the presence of bulky nodes larger than 2 cm was a criterion for excluding the patient from surgery. For limited disease, lymphadenectomy in these regions could be performed.

### Pathologic Evaluation

The protocol for pathologic evaluation of CRS specimens that was followed at the participating centres has been described in detail elsewhere.[20] This protocol was used for peritonectomy specimens for which no guidelines exist.

The primary ovarian tumour and lymph nodes were analysed and reported as per the existing guidelines.[21] Further details of pathological evaluation have been elaborated in a previous publication.

### Follow-up

Routine 3-monthly follow-up was performed for all patients with CA-125 levels and imaging studies as deemed suitable for the first two years and 6-monthly thereafter. The CA-125 level was checked after completion of the last cycle of chemotherapy and imaging studies performed if clinically indicated. Patients in whom the CA-125 returned to normal and those with no other clinical evidence of disease were considered to have had a complete response following first line therapy. The diagnosis of recurrence was made according to the Gynecologic Cancer Inter Group (GCIC) criteria.[22] The sites of recurrence were determined based on the first imaging study that showed evidence of recurrent disease.

Patients in whom the tumour markers failed to normalize after completion of first line therapy and/or had residual disease on imaging were considered to have platinum refractory disease. Platinum resistant recurrence (PRR) was defined as recurrence within 6 months of the last dose of platinum based therapy.

### Statistical Analysis

Categorical data are described as number (%). Abnormally distributed continuous data are expressed as median and range. Categorical data were compared with the v^2^ test. Median values were compared using the nonparametric independent sample t test, and means were compared using the Mann–Whitney U test. A p value lower than 0.05 was considered statistically significant.

Multivariable logistic regression was performed to evaluate the factors affecting grade 3-4 morbidity. Clinically relevant variables, as well as variables with a P value < 0.15 on univariate analysis, were evaluated in multivariable logistic regression model. The model was selected using a forward stepwise method. Odds ratios(OR) and their 95% confidence intervals (95%CI) were considered for logistic regression. Overall Survival (OS) was calculated from the date of surgery (iCRS+/-HIPEC) to the date of death or last follow up and progression-free survival (PFS) was determined from the date of surgery (iCRS +/- HIPEC) to the date of first recurrence. Survival curves were calculated using the Kaplan– Meier method and compared using the two-tailed log-rank test. Multivariable analyses using the Cox-proportional Hazard model, was used to calculate the factors affecting survival. Clinically relevant variables, as well as variables with a P value < 0.15 on univariate analysis, were entered for multivariable analyses. All statistical tests were two sided, with the significance level established at *p*-value of <0.05. All statistical analyses were conducted using SPSS version 22.0.0 (IBM Corporation, Armonk, NY, USA). Data collection was complete on 1^st^ May 2023.

## Results

From 9^th^ December 2018 to 31^st^ July 2022, 218 patients were enrolled at 4 Indian centres. The study flow-chart is provided in **Figure 1**. The median age 47 years [range 33-75] and 39(17.9%) patients were older than 65 years. 12 (5.5%) patients had low grade serous carcinoma, 5 (2.2%) had stage IV-A disease. 126 (57.7%) patients received 3 cycles of NACT and the remaining received more **(Table 1)**. One (0.4%) patient received neoadjuvant bevacizumab. BRCA testing was performed in 134 (61.4%) patients of which 12 (5.5%) had BRCA mutations. Homologous repair deficiency testing was not performed for any patient.

**Figure 1.**
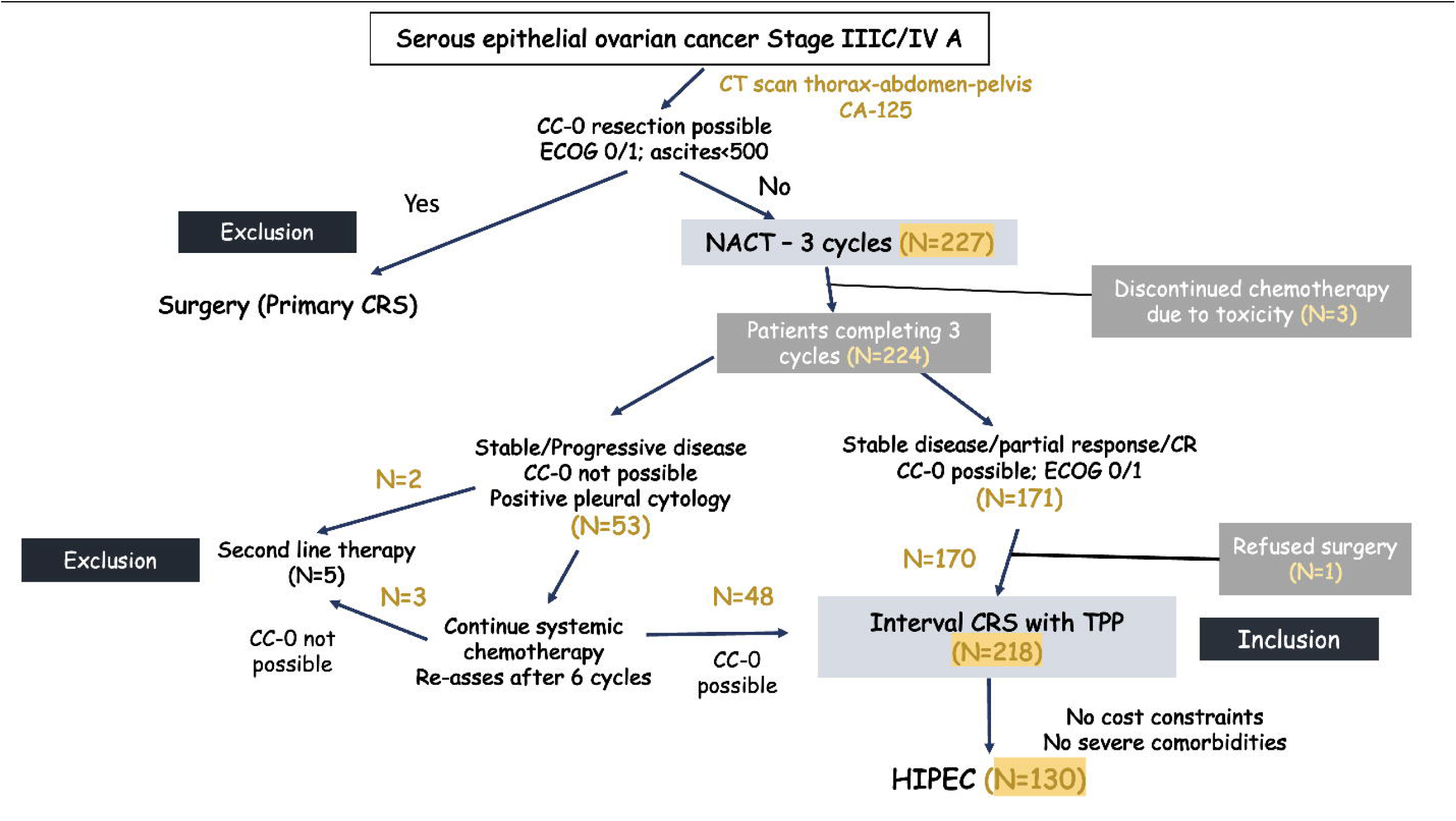
Study flow chart.

**Table 1.** Clinical and radiological features of 218 patient undergoing interval cytoreductive surgery with total parietal peritonectomy.

| Variable | All patients<br>N=218 (%) | iCRS+ HIPEC<br>N=130 (%) | iCRS<br>N=88 (%) | p-Value |
| --- | --- | --- | --- | --- |
| **Age** |  |  |  |  |
| ≤65 | 179 (82.1) | 116 (89.2) | 68 (77.2) | 0.016 |
| >65 | 39 (17.9) | 14 (10.8) | 20 (22.8) |  |
| **Histology** |  |  |  |  |
| Low grade serous | 12 (5.5) | 6 (4.6) | 6 (6.8) | 0.484 |
| High grade serous | 206 (94.5) | 124 (95.4) | 82 (93.2) |  |
| **FIGO Stage** |  |  |  |  |
| III-C | 213 (97.8) | 129 (99.2) | 85 (96.5) | 0.154 |
| IV-A | 5 (2.2) | 1 (0.8) | 3 (3.5) |  |
| **Cycles of NACT** |  |  |  |  |
| 3 | 126 (57.7) | 60 (46.1) | 66 (75.0) | <0.001 |
| 4 | 44 (20.1) | 37 (28.4) | 7 (7.9) |  |
| 5-6 | 48 (22.0) | 33 (25.3) | 15 (17.0) |  |
| **Pre-NACT CA-125 (IU/L)** |  |  |  |  |
| <500 | 51 (23.3) | 36 (27.6) | 15 (17.0) | 0.007 |
| 500-1000 | 33 (15.1) | 18 (13.8) | 15 (17.0) |  |
| >1000 | 70 (32.1) | 48 (36.9) | 22 (25.0) |  |
| Missing | 64 (29.3) | 28 (21.5) | 36 (40.9) |  |
| **Radiological PCI** |  |  |  |  |
| 0-10 | 85 (38.9) | 63 (48.4) | 22 (25.0) | 0.005 |
| 11-20 | 45 (20.6) | 40 (30.7) | 5 (5.6) |  |
| >20 | 6 (2.7) | 2 (1.5) | 4 (4.5) |  |
| Missing | 82 (37.6) | 25 (19.2) | 57 (64.7) |  |
| **Neoadjuvant bevacizumab** |  |  |  |  |
| Yes | 1 (0.4) | 0 (0.0) | 1 (1.1) | 0.780 |
| No | 217 (99.6) | 130 (100.0) | 87 (98.9) |  |
| **BRCA mutation** |  |  |  |  |
| Mutated | 12 (5.5) | 9 (6.9) | 3 (3.5) | 0.820 |
| HRD | No | No | No |  |
| Not mutated | 122 (55.0) | 95 (73.0) | 27 (30.6) |  |
| Not tested | 84 (39.5) | 26 (20.0) | 58 (65.9) |  |

The median surgical PCI was 14 [range 0-39]; 62.9% patients had PCI>10. A CC-0 resection was achieved in 167 (76.6%) and CC-1 in 42 (19.2%) patients. CC-1 was used when there were multiple nodules on the small bowel mesentery that were completely resected. So essentially, a CGR was achieved in 209 (95.8%) patients. HIPEC was performed in 130 (59.6%) patients. Details of the surgical procedures are provided in **Table 2**.

**Table 2.** Surgical findings and perioperative outcomes in 218 patients undergoing TPP during interval cytoreductive surgery.

| Variable | All patients<br>N=218 (%) | iCRS+ HIPEC<br>N=130 (%) | iCRS<br>N=88 (%) | p-Value |
| --- | --- | --- | --- | --- |
| **Surgical PCI** |  |  |  |  |
| 0-10 | 81 (37.1) | 56 (43.0) | 25 (28.4) | <0.001 |
| 11-20 | 90 (41.2) | 57 (43.8) | 33 (37.5) |  |
| 21-30 | 36 (16.5) | 17 (13.0) | 19 (21.5) |  |
| 31-39 | 11 (5.0) | 0 (0.0) | 11 (12.5) |  |
| **Median surgical PCI [range]** | 14 [0-39] | 12 [0-28] | 16 [1-39] |  |
| **CC-score** |  |  |  |  |
| CC-0 | 167 (76.6) | 110 (84.6) | 57 (64.7) | <0.001 |
| CC-1 | 42 (19.2) | 20 (15.3) | 22 (25.0) |  |
| CC-2/3 | 9 (4.12) | 0 (0.0) | 9 (10.2) |  |
| **Visceral resections** |  |  |  |  |
| Rectal resection | 87 (39.9) | 40 (30.7) | 47 (53.4) | <0.001 |
| Cholecystectomy | 126 (57.7) | 59 (45.3) | 67 (76.1) | <0.001 |
| Splenectomy | 58 (26.6) | 34 (26.1) | 24 (27.2) | 0.854 |
| Total colectomy | 9 (4.1) | 5 (3.8) | 4 (4.5) | 0.799 |
| Distal pancreatectomy | 2 (0.9) | 0 (0.0) | 2 (2.2) | 0.349 |
| Gastric resection | 1 (0.4) | 1 (0.8) | 0 (0.0) | 0.780 |
| Hepatic resection | 2 (0.9) | 1 (0.8) | 1 (1.1) | 0.780 |
| Appendicectomy | 136 (62.3) | 82 (63.0) | 54 (61.3) | 0.797 |
| **Lymphadenectomy** |  |  |  |  |
| Yes | 162 (74.3) | 97 (74.6) | 65 (73.8) | 0.900 |
| Pelvic | 162 (74.3) | 97 (74.6) | 65 (73.8) |  |
| Para-aortic | 90 (41.2) | 74 (56.9) |  |  |
| **Bowel anastomosis** |  |  |  |  |
| 0 | 114 (52.2) | 80 (61.5) | 34 (38.6) | 0.001 |
| 1 | 82 (37.6) | 37 (28.4) | 45 (51.1) |  |
| >1 | 22 (10.0) | 13 (10.0) | 9 (10.2) |  |
| **Visceral resections** |  |  |  |  |
| 0-3 | 105 (48.1) | 79 (60.7) | 26 (29.5) | <0.001 |
| >3 | 113 (51.9) | 51 (39.3) | 62 (70.5) |  |
| **Diaphragm resection** | 35 (16.0) | 18 (13.8) | 17 (19.3) | 0.280 |
| **Stoma** |  |  |  |  |
| Temporary | 19 (8.7) | 7 (5.3) | 12 (13.6) | 0.034 |
| Permanent | 0 | 0 (0.0) | 0 (0.0) |  |
| Mean operative time (mins) | 397.9 | 412.4 | 353.9 |  |
| Mean blood loss (ml) | 832.6 | 731.0 | 1032.5 |  |
| **Median ICU stay** | 2 [0-16] | 1 [0-12] | 3 [0-16] |  |
| **Median Hospital stay** | 10 [3-37] | 9 [3-37] | 14 [5-37] |  |
| **Grade 3-4 complications** | 38 (17.43) | 16 (12.3) | 22 (25.0) | 0.015 |
| **90-day mortality** | 6 (1.83) | 2 (1.5) | 4 (4.5) | 0.180 |
| **Time to adjuvant chemotherapy** |  |  |  |  |
| **<4 weeks** | 16 (7.3) | 15 (11.5) | 1 (1.1) | 0.037 |
| **4-6 weeks** | 162 (74.3) | 90 (69.2) | 72 (81.8) |  |
| **>6 weeks** | 16 (7.3) | 8 (6.1) | 8 (9.0) |  |
| **Planned but not started** | 2 (0.9) | 2 (1.5) | 0 (0.0) |  |
| **Not planned** | 22 (10.0) | 15 (11.5) | 7 (7.9) |  |

### Variation in the clinical and surgical findings at the 4 centres

One centre enrolled less than 30 patients while the other three enrolled more than 60 patients each **(Table 3)**. The median surgical PCI was significantly higher at centres 1 and 2 compared to centres 3 and 4 and the average no of visceral resections was also significantly higher at these two centres. Contrary to this significantly fewer patients received HIPEC at centres 1 and 2 compared to centres 3 and 4.

**Table 3.** Variation in demographic, clinical, surgical and pathological findings according to the centre.

| Variable | All patients | Centre 1<br>(Zydus) | Centre 2<br>(Saifee) | Centre 3<br>(Jehangir) | Centre 4<br>(MVR) | p-value |
| --- | --- | --- | --- | --- | --- | --- |
| Recruitment commencement | 2018-2022 | 2018-2022 | 2018-2022 | 2018-2022 | 2021-2022 |  |
| No of patients | 218 | 62 | 69 | 26 | 61 |  |
| NACT cycles |  |  |  |  |  |  |
| 3 | 126 (57.7) | 46 (74.1) | 41 (59.4) | 19 (73.0) | 20 (32.7) | <0.001 |
| 4 | 44 (20.1) | 6 (9.6) | 11 (15.9) | 1 (3.8) | 26 (42.6) |  |
| 5-6 | 48 (22.0) | 10 (16.1) | 17 (24.6) | 6 (23.0) | 15 (24.5) |  |
| Median surgical PCI [range] | 14 [0-39] | 16 [2-37] | 17 [0-39] | 13 [1-26] | 8 [2-20] | <0.001 |
| HIPEC | 129 (59.1) | 26 (41.9) | 20 (28.9) | 23 (88.4) | 60 (98.3) | <0.001 |
| No of bowel anastomosis |  |  |  |  |  |  |
| 0 | 114 (52.2) | 11 (17.7) | 32 (46.3) | 20 (76.9) | 51 (83.6) | <0.001 |
| 1 | 82 (37.6) | 41 (66.1) | 26 (37.6) | 6 (23.1) | 9 (14.7) |  |
| >1 | 22 (10.0) | 10 (16.1) | 11 (15.9) | 0 (0.0) | 1 (1.6) |  |
| Visceral resections |  |  |  |  |  |  |
| <3 | 104 (47.8) | 20 (35.4) | 12 (17.3) | 22 (84.6) | 50 (81.9) | <0.001 |
| >3 | 114 (52.2) | 42 (64.6) | 57 (82.7) | 4 (15.4) | 11 (18.1) |  |
| Lymphadenectomy | 162 (74.3) | 51 (82.2) | 32 (46.3) | 20 (76.9) | 59 (96.7) | <0.001 |
| Rectal resection | 87 | 41 (66.1) | 35 (50.7) | 5 (19.2) | 6 (9.8) | <0.001 |
| Cholecystectomy | 124 (56.8) | 55 (88.7) | 55 (79.7) | 7 (26.9) | 7 (11.4) | <0.001 |
| Splenectomy | 58 (26.6) | 29 (46.7) | 17 (24.6) | 8 (30.7) | 4 (6.5) | <0.001 |
| Grade 3-4 complications | 38 (17.4) | 20 (32.2) | 13 (18.8) | 3 (11.5) | 2 (3.2) | <0.001 |
| 90-day mortality | 6 (2.7) | 3 (4.8) | 1 (1.4) | 1 (3.8) | 1 (1.6) | 0.605 |
| Pathological PCI |  | 8 [0-36] | 13 [0-25] | 11 [0-26] | 6 [0-15] |  |
| Chemo response score |  |  |  |  |  |  |
| 1-2 | 174 (79.8) | 55 (88.7) | 50 (72.4) | 22 (84.6) | 47 (77.0) | 0.557 |
| 3 | 26 (11.9) | 7 (11.3) | 5 (7.2) | 4 (15.4) | 10 (16.3) |  |
| Missing | 28 (12.8) | 0 (0.0) | 14 (20.2) | 0 (0.0) | 4 (6.5) |  |
| Upper abdomen involvement | 145 (66.5) | 40 (64.5) | 57 (82.6) | 17 (65.3) | 31 (50.8) | <0.001 |
| Lymph node involvement | 63 (28.8) | 20 (32.2) | 17 (24.6) | 9 (34.6) | 19 (31.1) | 0.705 |
| Median follow-up (months) | 18 | 34 | 19 | 21 | 12.5 | <0.001 |
| No of recurrences | 97 | 37 | 27 | 12 | 21 | 0.028 |
| Progression-free survival | 20 | 19 | 26 | 25 | 19 | 0.101 |
| Early recurrence | 41 | 18 | 6 | 7 | 10 | 0.042 |
| Platinum-resistant recurrence | 15 | 5 | 3 | 3 | 4 | 0.094 |
| Secondary CRS | 11 | 6 | 5 | 0 | 0 | 0.445 |

### 90-day morbidity and mortality

Grade1 and 2 complications occurred in 27 (12.3%) and grade 3 and 4 complications in 38 (17.4%) patients within 90 days of surgery **(Table 4).** The most common complication was post-op intra-abdominal fluid collection requiring drainage that was observed in 15 (6.8%), followed by systemic sepsis in 9 (4.1%) and post-operative haemorrhage in 6 (2.7%) patients. Six (2.7%) patients died within 90-days of surgery of which 2 death were due to post- operative complications (one due to haemorrhagic shock and 1 due to systemic sepsis). Two patients died of myocardial infarction, one of COVID-19 infection and one due to chemotherapy related complications. The major morbidity was significantly higher at the first two centres but the 90-day mortality was similar at all 4 centres **(Table 3)**. Adjuvant chemotherapy was planned for 196 patients of which 194 (99.1%) started the treatment. 7.3% started adjuvant therapy within 4 weeks of surgery; 74.3% in 4-6 weeks and in 7.3%, it was delayed beyond 6 weeks. The factors affecting grade 3-4 morbidity are described in **Table 5**. On multivariate analysis, the centre was the only factor affecting major morbidity

**Table 4.** Factors affecting progression-free survival.

| Variable | Progression-free survival |  |  | Overall survival |  |  |
| --- | --- | --- | --- | --- | --- | --- |
|  | Univariate analysis<br>p-value | Multivariate analysis |  | Univariate analysis<br>p-value | Multivariate analysis |  |
|  |  | Hazard ratio | p-value |  | Hazard ratio | p-value |
| Age <45 vs >45 | 0.089 |  |  | 0.253 |  |  |
| Center | 0.101 |  |  | 0.659 |  |  |
| Surgical PCI | 0.374 |  |  | 0.072 |  |  |
| Surgical PCI <10 vs >10 | 0.110 |  |  | 0.022 |  |  |
| Surgical PCI <15 vs >15 | 0.035 |  |  | 0.015 |  |  |
| CC-0 vs CC 1-3 | 0.078 |  |  | 0.317 |  |  |
| Path PCI <15 vs >15 | 0.019 |  |  | 0.007 |  |  |
| Path PCI <10 vs >10 | 0.112 |  |  | 0.225 |  |  |
| Path PCI | <0.001 |  | 0.028 | <0.001 |  | 0.006 |
| NACT 3 cycles vs >3 cycles | 0.080 |  |  | 0.753 |  |  |
| Grade 3-4 complications | 0.740 |  |  | 0.915 |  |  |
| Time to adjuvant chemotherapy | 0.034 |  |  | 0.021 |  | 0.004 |
| HIPEC | 0.272 |  |  | 0.147 |  |  |
| BRCA status | 0.025 |  |  | 0.935 |  |  |
| Disease in normal peritoneum | 0.105 |  |  | 0.232 |  |  |
| Positive lymph nodes | 0.469 |  |  | 0.972 |  |  |
| Disease in upper regions | 0.101 |  |  | 0.176 |  |  |
| Disease on small bowel or its mesentery | 0.073 |  |  | 0.001 |  | 0.001 |
| Splenectomy (no vs yes) | 0.598 |  |  | 0.029 |  |  |
| Bowel resection | 0.886 |  |  | 0.570 |  |  |
| Bowel anastomoses | 0.138 |  |  | 0.239 |  |  |
| Organ resections 0-3 vs >3 | 0.314 |  |  | 0.294 |  |  |
| Duration of surgery <8 hours | 0.077 |  |  | 0.080 |  |  |
| Rectosigmoid resection | 0.719 |  |  | 0.306 |  |  |

**Table 5.** Complications occurring in patients treated with and without HIPEC.

|  | All patients<br>N=218 (%) | HIPEC<br>N=130 (%) | No HIPEC<br>N=88 (%) | p-Value |
| --- | --- | --- | --- | --- |
| Grade 1-2 complications | 27 (12.3%) | 13 (10.0) | 14 (15.9%) |  |
| **Grade 3-4 complications** | 38 (17.4) | 16 (12.3) | 22 (25.0) | 0.015 |
| **90-day mortality** | 6 (2.7) | 2 (1.5) | 6 (6.8) | 0.180 |
| Type of complications |  |  |  |  |
| Haemorrhage | 6 (2.7) | 4 (3.0) | 2 (2.2) | 0.721 |
| Hematologic | 1 (0.4) | 1 (0.8) | 0 (0.0) | 0.780 |
| Respiratory | 3 (1.3) | 3 (2.3) | 2 (2.2) | 0.986 |
| Sepsis | 9 (4.1) | 6 (4.6) | 3 (3.4) | 0.660 |
| Neutropenia | 0 (0.0) | 0 (0.0) | 0 (0.0) | . |
| Cardiac | 2 (0.9) | 0 (0.0) | 2 (2.2) | 0.349 |
| Gastrointestinal | 5 (2.2) | 3 (2.3) | 2 (2.2) | 0.986 |
| Bowel fistula | 4 (1.8) | 2 (1.5) | 2 (2.2) | 0.691 |
| Bowel perforation | 0 (0.0) | 0 (0.0) | 0 (0.0) | . |
| Nephrological | 1 (0.4) | 1 (0.8) | 0 (0.0) | 0.780 |
| Urological | 1 (0.4) | 1 (0.8) | 0 (0.0) | 0.780 |
| Wound dehiscence | 2 (0.9) | 0 (0.0) | 2 (2.2) | 0.691 |
| Surgical site infection | 0 (0.0) | 0 (0.0) | 0 (0.0) | . |
| Post op intra-abdominal fluid collection | 15 (6.8) | 6 (4.6) | 9 (10.2) | 0.108 |
| Return to operating room | 10 (4.5%) | 6 (4.6) | 4 (4.5) | 0.980 |

### Pathological findings

The median pathological PCI was 8 [range 0-36]. Involvement of normal appearing peritoneum was observed in 77 (35.3%) and involvement of peritoneum around tumour nodules in 48 (22.0%) patients **(Table 6).** 29 (13.3%) patients had a Bohm’s score of 3 (complete or near complete response).

**Table 6.** Factors affecting grade 3-4 morbidity.

| Clinical variable | Univariate analysis | Multivariate analysis |
| --- | --- | --- |
| Age (<45 versus >45) | 0.246 |  |
| Surgical PCI (<10 versus >10) | 0.017 | 0.182 |
| Surgical PCI (<15 versus >15) | 0.044 | 0.867 |
| No of NACT cycles (3 versus >3) | 0.030 | 0.051 |
| CC-0/1 versus CC 2/3 | 0.054 | 0.932 |
| HIPEC (yes versus no) | 0.077 | 0.196 |
| Pathological PCI <15 versus >15 | 0.215 |  |
| **Centre** | **<0.001** | **0.002** |
| Disease in upper regions (1-3) | 0.672 |  |
| Splenectomy | 0.095 |  |
| Small bowel resection | 0.011 | 0.221 |
| Visceral resections (<4 versus 4 or more) | 0.020 | 0.874 |
| Duration of surgery (<480 versus >480 mins) | 0.196 |  |
| Rectosigmoid resection (yes versus no) | <0.001 | 0.341 |

### Platinum resistant recurrence and survival outcomes

At a median follow-up 19 months [95% confidence interval (CI) 15-35months], 101 (46.3%) patients had recurred and 19 (8.7%) patients were dead. All patients had at least 6 months of follow-up from the date of surgery or last cycles of platinum-based chemotherapy whichever was last. Platinum refractory disease was seen in 9(4.1%) and PRR in 15 (6.8%) patients. The median PFS was 22 months [95% CI 17-35months]. The median OS was not reached. The projected 3-year OS was 81.5% and in 80 patients who were treated before May 2020, it was 77.5%. Due to the small no of patients with PRR, factors affecting it could not be studied. The univariate and multivariate analyses of factors affecting PFS are presented in **Table 6**. The pathological PCI was the only factor independently influencing PFS.

## Discussion

This study shows that TPP during iCRS can be performed with an acceptable grade 3-4 morbidity (17.4%) and post-operative mortality (2.7%).[17,23] This confirms our previous findings in the first 50 patients enrolled in the study.[14] The incidence of PRR was low (6.8%) and similar to what we have reported previously.[15,24]

The overall incidence of potentially life-threatening complications like bowel fistula and perforation was low **(Table 5)**. The major morbidity was significantly higher at two centres which could be attributed to a higher median surgical PCI and more extensive surgery performed at these centres. However, the 90-day mortality was similar among all centres **(Table 3)**. There are several reasons for the variation in the extent of surgery between the different centres. The referral pattern at the centres was different. The first two surgeons were likely to receive referrals of patients with more extensive disease as reflected in the median surgical PCI. At one centre fewer patients received surgery after 3 cycles of NACT compared to the other 3 centres. This may reflect a logistic issue or surgeon’s preference as it was not mandatory in the study to operate after 3 cycles of NACT. Third, though the surgical protocol was fixed, the visceral resection cannot be pre-specified and will depend both on intraoperative findings and the surgeons approach towards resections of scars and minimal residual disease. We consider this to be an important finding from this study that highlights the how iCRS can vary in extent. This difference can also be observed in two recent randomized trials. In the KOVHIPEC-1 trial, more than 80% patients undergoing iCRS had diaphragmatic stripping and more than 60% had a rectal resection while in the SCORPION trial, these figures were less than 30% and less than 20% respectively.[3,23] The final survival outcomes will indicate if a more aggressive approach to visceral resection yields a survival benefit or not.

The incidence of respiratory complications was low considering that all patients had bilateral diaphragmatic stripping. Patients at all 4 centres are made to do physical exercise and respiratory exercises on the day of the first consultation with the surgical team itself. The prehabilitation combined with the selective use of non-invasive ventilation after cessation of invasive ventilation has greatly reduced the respiratory complications in our experience.[14,25] The two concerning complications are haemorrhage and systemic sepsis. Though the incidence of both was less than 10%, all patients with post operative haemorrhage required reoperation. Haemorrhage is less common after Cisplatin HIPEC and we attribute it to the extensive surgery rather than HIPEC. HIPEC itself was not associated with an increased incidence of complications. On the contrary, patients who did not undergo HIPEC had a significantly higher incidence of complications which could be attributed to the more extensive surgery in that group. Systemic sepsis remains a country specific problem as we have highlighted in our previous reports.[25,26] The incidence of renal complications was low. Sodium thiosulfate is not available in India and therefore, we used a lower dose of Cisplatin which was shown to be safe in a phase 1 trial.[27,28] Pre-hydration with intravenous fluids for at least 12 hours prior to surgery was used at all 4 centres in patients who were likely to undergo HIPEC.

Though we did not define the upper limit of age for inclusion in the study, all 4 surgeons were more cautious in including patients aged over 65 years in this study. This is reflected in the fact that less than 20% of the patients were aged over 65 years. In our experience, the tolerance of older Indian patients to such extensive surgery is less than that reported by European and North American centres where the patient population is predominantly Caucasian or that reported from South Korea. In the KOVHIPEC-1 trial, the median time to adjuvant chemotherapy was 21 days while in the current study, only 7.3% of the patients were able to start adjuvant chemotherapy within four weeks of surgery.[23] Our target for starting adjuvant chemotherapy is within 6 weeks as a delay of more than 6 weeks has demonstrated a negative impact on survival.

One of the biggest strengths of this study is that the 90-day morbidity was recorded. It has been shown that complications can occur after discharge and up to 90-days of CRS and HIPEC.[3,29] There were two deaths due to myocardial infarction. This could be considered an indirect consequence of surgery where the capacity to tolerate physical stress is reduced. Both deaths occurred in patients aged more than 65 years.

BRCA testing was performed only in 61% as only those patients who can afford PARP inhibitors undergo BRCA testing. The cost of 5 months of Olaparib is more than the cost of iCRS with HIPEC in our set-up. With the recent results of the Athena mono trial, the BRCA and HRD testing has increased.[30] Though the incidence of BRCA mutations is low, we will analyse and report survival outcomes in this subgroup in the future. We use PFS as a primary end-point as it is used in many major randomized trials on ovarian cancer. It is also an end- point that is not influenced by the subsequent systemic treatments that may undermine the benefit of the treatment in question. PFS is important to patients as it represents being cancer-free and many patients will be treatment-free as well.[31]

The PRR remains low as reported previously. This provides further proof for our hypothesis that removing previously involved peritoneum could delay recurrence if not prevent it. The PFS and OS data are not mature but 101 (46.3%) recurrences have occurred and the median PFS is 22 months which is encouraging.

The main limitation of this study is the variation in the extent of visceral resections. Though we had a fixed surgical protocol, in the setting of peritoneal disease, it is difficult to predefine the extent of surgery completely. We did not use predictive scores for selecting patients for pCRS versus iCRS but a careful evaluation of the prechemotherapy scans was done to ensure that only patients with stage III-C and above receiving NACT were included in the study. There were several reasons for this- many times patients are referred directly after NACT and this study captures our day-to-day practice scenario; staging laparoscopy before NACT is required for calculating predictive scores which is not feasible in many patients due to the incremental cost; in addition to the feasibility of a complete resection, the age and performance status are very important for us in selecting patients for pCRS versus iCRS. The strengths are the study design itself that represents a meticulous approach to addressing the parietal peritoneum, the most common site of residual disease and capturing of the 90-day morbidity and mortality.

## Conclusions

The 90-day grade 3-4 morbidity and mortality of TPP with or without HIPEC performed during iCRS is acceptable. It was the extent of surgery and not HIPEC that had an impact on the morbidity. The incidence was of PRR is low. Early survival results are encouraging and warrant conduction of a randomized controlled trial comparing TPP with conventional surgery.

## Data Availability

Data will be made available upon reasonable request to the corresponding author

https://pubmed.ncbi.nlm.nih.gov/32748154/

## Conflict of interests

The authors have no disclosures

The author report no conflict of interests

## Notes

**Disclosures and conflicts of interest** The authors have no disclosures The authors report no conflict of interests

### Competing Interest Statement

The authors have declared no competing interest.

### Clinical Trial

CTRI/2018/12/016789 This study is registered with the clinical trials registry of India

### Clinical Protocols

https://www.ctri.nic.in

https://www.ctri.nic.in/

### Funding Statement

This study did not receive any funding

### Author Declarations

Zydus Hospital Ethics Committee approved this study on 8th December 2018 Address: Zydus hospital, Zydus hospital road, Ahmedabad, 380054, India

