## Supplement 1 for "Perioperative outcomes and platinum resistant recurrence in patients undergoing protocol based total parietal peritonectomy during interval cytoreductive surgery for advanced ovarian cancer- results of the TORPEDO study"

**Surgical protocol**

**Total parietal peritonectomy (TPP)**

TPP comprises of all 5 peritonectomies- that is pelvic, bilateral anteroparietal, right and left upper quadrant peritonectomies with a total omentectomy. In case of TPP, all the regions marked in bold have to be removed irrespective of tumor involvement or not.

Exceptions include removal of the Glisson’s capsule in regions 1 and 2 in absence of tumor and the superior and inferior recesses of the lesser sac.

For mesenteric disease, performance of mesenteric peritonectomy is at the discretion of the surgeon. A minimal of representative biopsy of tumor bearing or normal mesenteric peritoneum (if no disease is present) should be performed separately for each of the regions from 9-12

**Algorithm**

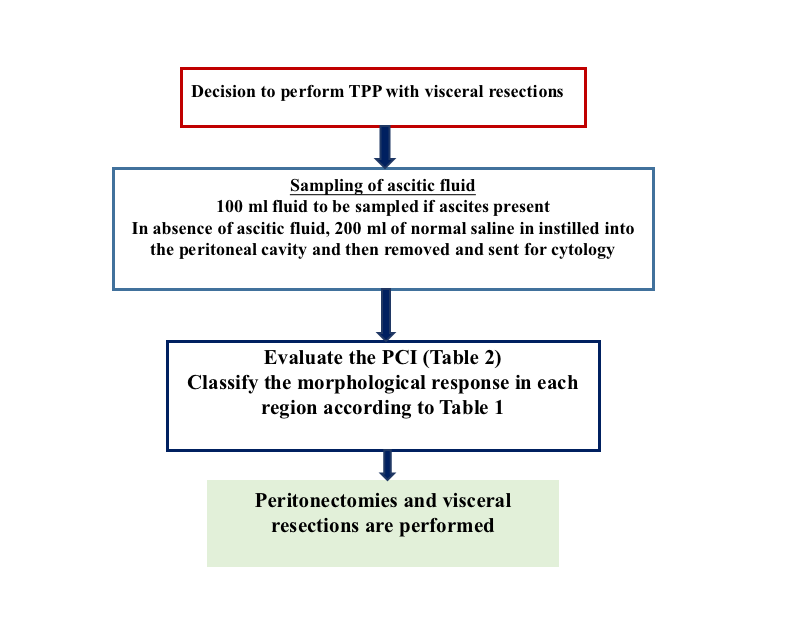

**Table 1 Visual assessment of presence or absence of tumor**

| **Positive for tumor** | **Negative for tumor** |
| --- | --- |
| Tumor nodule/s  Confluent deposit  Omental cake  Plaque  Thickening  Scarring | Thickening  Scarring  None of the above |

**Table 2- detailed surgical PCI and evaluation of tumor in each region**.

The regions in bold are the parietal peritoneal regions. When tumor is present, select one option from the 4^th^ column and if tumor is absent select one option from the 5^th^ column

| Region | Structure | Size of largest deposit | Nodule (N)  Plaque (P)  Confluent (CO)  Cake (CK)  Thickening (T)  Scarring (S) | Thickening  Scarring  No tumor | Size of residual disease | Score (for the entire region) |
| --- | --- | --- | --- | --- | --- | --- |
| 0 | **Midline incision (scar if removed)** |  |  |  |  |  |
|  | **Anteroparietal peritoneum (Right)** |  |  |  |  |  |
|  | **Anteroparietal peritoneum (Left)** |  |  |  |  |  |
|  | **Greater omentum** |  |  |  |  |  |
|  | **Inferior recess of lesser sac** |  |  |  |  |  |
|  | **Remnant greater omentum** |  |  |  |  |  |
|  | Transverse colon |  |  |  |  |  |
|  | Transverse mesocolon |  |  |  |  |  |
| 1 | Right Glisson’s |  |  |  |  |  |
|  | Right inferior Glisson’s |  |  |  |  |  |
|  | **Right subphrenic peritoneum** |  |  |  |  |  |
|  | **Morrison’s pouch** |  |  |  |  |  |
|  | Gall bladder |  |  |  |  |  |
| 2 | Left Glisson’s |  |  |  |  |  |
|  | **Lesser omentum** |  |  |  |  |  |
|  | **Hepatoduodenal ligament** |  |  |  |  |  |
|  | **Left central region** |  |  |  |  |  |
|  | **Falciform ligament** |  |  |  |  |  |
|  | **Tissue in umbilical fissure** |  |  |  |  |  |
|  | Lesser sac superior recess |  |  |  |  |  |
|  | **Lesser sac inferior recess** |  |  |  |  |  |
| 3 | **Left diaphragmatic peritoneum** |  |  |  |  |  |
|  | Splenic surface |  |  |  |  |  |
|  | Tail of pancreas/splenic hilum |  |  |  |  |  |
|  | Anterior surface of the stomach |  |  |  |  |  |
|  | Posterior surface of the stomach |  |  |  |  |  |
| 4 | **Left paracolic peritoneum** |  |  |  |  |  |
|  | Left mesocolon |  |  |  |  |  |
|  | Left colon |  |  |  |  |  |
|  | Splenic flexure of colon |  |  |  |  |  |
| 5 | **Left pelvic peritoneum** |  |  |  |  |  |
|  | Sigmoid colon |  |  |  |  |  |
|  | Sigmoid mesocolon |  |  |  |  |  |
|  | Appendices epiploicae |  |  |  |  |  |
| 6 | **Bladder** |  |  |  |  |  |
|  | **Pouch of Douglas** |  |  |  |  |  |
|  | **Right pararectal peritoneum** |  |  |  |  |  |
|  | **Left pararectal peritoneum** |  |  |  |  |  |
|  | Left fallopian tube |  |  |  |  |  |
|  | Left ovary |  |  |  |  |  |
|  | Right fallopian tube |  |  |  |  |  |
|  | Right ovary |  |  |  |  |  |
|  | Uterus |  |  |  |  |  |
|  | Rectosigmoid junction |  |  |  |  |  |
| 7 | **Right pelvic peritoneum** |  |  |  |  |  |
|  | Caecum |  |  |  |  |  |
|  | Appendix |  |  |  |  |  |
|  | Mesoappendix |  |  |  |  |  |
| 8 | **Right paracolic peritoneum** |  |  |  |  |  |
|  | Right colon |  |  |  |  |  |
|  | Right mesocolon |  |  |  |  |  |
|  | Appendices epliplicae |  |  |  |  |  |
| 9 | Proximal jejunal surface |  |  |  |  |  |
|  | Proximal jejunal surface (mesenteric) |  |  |  |  |  |
|  | Proximal mesojejunum |  |  |  |  |  |
| 10 | Distal jejunal surface |  |  |  |  |  |
|  | Distal jejunal surface (mesenteric) |  |  |  |  |  |
|  | Distal mesojejunum |  |  |  |  |  |
| 11 | Proximal ileal surface |  |  |  |  |  |
|  | Proximal ileal surface (mesenteric) |  |  |  |  |  |
|  | Proximal mesoileum |  |  |  |  |  |
| 12 | Distal ileal surface |  |  |  |  |  |
|  | Distal ileal surface (mesenteric) |  |  |  |  |  |
|  | Distal mesoileum |  |  |  |  |  |
| Total | | | | |  |  |

**Guidelines for peritonectomy procedures**

A midline incision from the xiphoid to the pubis is to be employed for all cases irrespective of the disease extent.

- Scars of previous CRS or diagnostic procedures to be excised.
- Drain and port sites to be excised. Scars of non- cancer surgery may not be excised.
- Approach could be extra peritoneal or intra peritoneal provided the evaluation of the PCI is accurate.

1. **Pelvic peritoneal regions**

**Right pelvic peritoneum**- begins at the root of the appendix and lower border of the caecum, including the lower anterior parietal peritoneum and the peritoneum over the psoas and external iliac vessels. The medial boundary is formed by the ureter. Inferiorly it extend till the lateral end of the bladder peritoneum.

**Left pelvic peritoneum-** upper limit at the lateral border of the descending sigmoid junction and follows the lateral edge of the sigmoid. The other limits correspond to those on the right.

**Bladder peritoneum**- peritoneum overlying the urinary bladder till the vesicovaginal fold inferiorly. Laterally, it is bounded by the lower end of the broad ligament in females. In males the inferior boundary is the pouch of Douglas.

**Right pararectal peritoneum**- from the sacral promontory superiorly, to the POD inferiorly, the ureter laterally to the attachment on the rectal wall medially.

**Left pararectal peritoneum**- similarly on the right side.

**Root of the sigmoid**- bounded by the origin of the inferior mesenteric artery superiorly, the sigmoid to the left, the root of the appendix and caecum to the right and the peritoneum overlying the sacral promontory inferiorly

2. **Anteroparietal peritonectomies** (right and left)

From the midline incision anteromedially to the line of Todt posteromedially. It merges inferiorly on the right side with the right pelvic peritoneum and on the left side with the left pelvic peritoneum.

Divided into the paracolic peritoneum (right and left) that extends from the line of Todt to the lateral boundary of the paracolic gutter.

The rest of the peritoneum in the region on each side forms the anteroparietal peritoneum.

The upper limit is at the inferior pole of the kidney on each side.

3. **Right upper quadrant regions**

**Right subphrenic peritoneum**- From the right of the falciform ligament medially it includes all the peritoneum on the undersurface of the right dome, extending inferiorly to the lower pole of the right kidney. Postero-superiorly, it merges with the glisson’s capsule at the superior boundary of the right lobe. The coronary ligament has to be divided completely to excise this part of the peritoneum. Postero-medially it is attached to the posteromedial edge of segment 6 and 7. Division of this attachment exposes the retrohepatic inferior vena cava. Medially it follows the inferior edge of segments 5 and 6 to the lateral edge of the porta hepatis and includes the peritoneum on the second and third parts of the duodenum.

**Morrisons’ pouch**- Laterally from the medial boundary of the right kidney, superiorly, the inferior surface of segments 5 and 8, medially the lateral edge of the hepatoduodenal ligament and inferiorly, the superior and lateral edge of the duodenum and inferolaterally, the hepatic flexure.

**Right glisson’s**- Capsule of the right lobe of the liver, superior and lateral surfaces. Extends from the right side of the attachement of the falciform onto the superior surface of the right lobe and the lateral surface, merging with the attachment of the right subphrenic peritoneum in that region.

**Right inferior glisson’s**- the part of the capsule on the inferior surface of the right lobe and the right caudate lobe.

4. **Left upper quadrant peritoneal regions**

**Falciform ligament**- from the umbilicus it extends along the inferior surface of the anterior abdominal wall till diaphragm. Inferior attachement on the superior surface of the liver. Inferiorly, it forms the umbilical ligament in the umbilical fissure containing the obliterated umbilical vein.

**Tissue in the umbilical fissure**- removed separately. Superiorly, it merges with the falciform ligament and inferiorly with the hepatoduodenal ligament

**Left central peritoneum**- From the left edge of the falciform ligament till the left lateral boundary of the esophageal hiatus and interiorly it merges with the peritoneal reflection on the superior edge of the left lobe of the liver. The left upper antero parietal peritoneum is not included in this region.

**Left subphrenic peritoneum**- Extends from the midline incision laterally to include all the peritoneum on the undersuface of the left dome of the diaphragm merging postero inferiorly with the upper end of the left antero parietal peritoneum, superomedially the lateral edge of the abdominal esophagus and fundus, and along the lateral edge of the spleen inferomedially. Removal of the peritoneum from the spleen should extend right upto the hilum posteriorly and expose the splenic hilar vessels.

**Left Glisson’s capsule**- Capsule on the superior and inferolateral surface of the left lobe, to the left of the falciform ligament

5. **Total omentectomy regions**

**Hepatoduodenal ligament**- peritoneum overlying the porta hepatis extending from the lateral border of the common bile duct to the medial border of the portal vein and the lower end of the umbilical ligament to the superior border of the first part of the duodenum

**Lesser omentum** extends from the medial boundary of the hepatoduodenal ligament inferiorly to the left coronary ligament superiorly, inferomedially it is attached to the lesser curve and superiorly to the inferior surface of the left lobe overhanging the caudate. The left coronary ligament needs to be divided completely to excise the lesser omentum completely. The gastric arcade is preserved in absence of gross tumor deposits in this region

**Lesser sac superior recess**- The caudate lobe is reflected laterally and the peritoneum over the vena cava and the right crus is excised completely

**Lesser sac inferior recess**- includes the anterior leaf of the transverse mesocolon, the pancreatic capsule and the gastro pancreatic fold of peritoneum. The capsule over entire head, body and tail of the pancreas is removed.

**Infracolic greater omentum**

Greater omentectomy begins along the line of Todt, resecting the omentum over the ascending and descending colon on the right and left sides respectively, on the right it includes the part over the hepatic flexure and the peritoneum over the c-loop of the duodenum; on the left it is divided at its attachment to the inferior pole of the spleen and the inferomedial surface. The omentum may be divided along its attachment to the transverse colon to perform an infracolic omentectomy alone.

**Supracolic omentectomy**- The division the the omentum along to transverse colon is performed in a manner to open up the plane between the anterior and posterior layers of the transverse mesocolon and the anterior layer is divided at its attachment to the inferior border of the pancreas or the pancreatic capsule is resected in continuity. If the pancreatic capsule is not resected, the dissection reaches the greater curve. The gastro omental vein is divided. The arc of Barkov is preserved in absence of gross disease in the omentum and removed in case of gross disease.

**Guidelines for visceral resections**

Any visceral that is infiltrated by the tumor needs to be resected to obtain a complete cytoreduction. Visceral sites showing a complete clinical response need not be resected like the parietal peritoneum.

**Resection of the uterus, tubes and ovaries-** It is to be performed for all patients. The infundibulopelvic ligaments are divided as high as possible. These structures are usually resected en-bloc with a pelvic peritonectomy. If a hysterectomy has been performed below, the peritoneum overlying the vaginal stump should be removed. The vaginal stump may be opened and resutured in case of adherent deposits.

**Resection of the rectum-** Pelvic peritoneal deposits infiltrating the rectum require resection of the rectum. In case, there is a good response, the rectal resection may be omitted but the pelvic peritoneum must be completely stripped off the rectum. When a rectal resection is performed a total mesorectal excision akin to that for primary rectal carcinomas should be performed. The decision to perform a diverting stoma is at the discretion of the operating surgeon but for all anastomosis below the level of the peritoneal reflection, a diverting stoma is recommended.

**Resection of the sigmoid**- It may be required in case of gross disease on the serosa or due to multiple deposits on the appendices epiploicae and the mesentery which cannot be resected individually.

**Cholecystectomy** – It is to be performed for all patients irrespective of the presence or absence of disease

**Splenectomy** – It is to be performed in case or residual disease. It can be preserved if there are few nodules that can be completely excised and are not infiltrating the parenchyma.

Appendectomy- May not be performed for all patients unless there is presence of disease. No mucinous tumors are included in this study and hence an appendectomy is not indicated in absence of disease.

**Resection of other viscera**- resection of the liver segments, pancreas, small bowel and colon is performed in there are infiltrative deposits that require a resection of the organ for obtaining a complete cytoreduction.

Resection of bladder, ureters and/or kidney may be performed to obtain a complete cytoreduction if required.

**Guidelines of lymphadenctomy**

A bilateral pelvic and retroperitoneal lymphadenectomy is performed for all patients in whom there is suspicion of nodal disease on imaging or during surgery. The upper extent is at the level of the renal veins for a retroperitoneal lymphadenectomy. In case of suprarenal lymphadenopathy, presence of bulky nodes >2cm would exclude the patient from surgery. For limited disease, lymphadenectomy in these regions could be performed.

The lymph nodes removed along with bowel segments, peritoneal fat and omentum are to be analyzed according to the guidelines for pathological evaluation.
